# Impact of COVID-19 on Migrants’ Access to Primary Care: A National Qualitative Study

**DOI:** 10.1101/2021.01.12.21249692

**Authors:** Felicity Knights, Jessica Carter, Anna Deal, Alison F Crawshaw, Sally E Hayward, Lucinda Jones, Sally Hargreaves

## Abstract

**Background:** The COVID-19 pandemic has led to considerable changes in the delivery of primary care in the UK, including rapid digitalisation, yet the extent to which these have impacted on marginalised migrant groups – already facing existing barriers to NHS care – is unknown. Understanding the perspectives and experiences of health professionals and migrants will support initiatives to deliver more effective health services, including delivery of the COVID-19 vaccine, to marginalised groups.

**Aim:** To understand the impact of the COVID-19 pandemic on migrants and their access to primary healthcare, and implications for COVID-19 vaccine roll out.

**Design and Setting:** Primary care professionals, administrative staff, and migrants (foreign born; >18 years; <10 years in UK), were recruited in three phases using purposive, convenience and snowball sampling from urban, suburban and rural settings.

**Methods:** In-depth semi-structured interviews were conducted by telephone. Data were analysed iteratively, informed by thematic analysis.

**Results:** 64 clinicians were recruited in Phase 1 (25 GPs, 15 nurses, 7 HCAs, 1 Pharmacists); Phase 2 comprised administrative staff (11 PMs and 5 receptionists); and in Phase 3 we recruited 17 migrants (88% asylum seekers; 65% female; mean time in UK 4 years). We found that digitalisation and virtual consultations (telephone, video, and online form-based) have amplified existing inequalities in access to healthcare for many migrants due to lack of digital literacy and access to technology, compounded by language barriers. Use of virtual consultations has resulted in concerns around building trust and the risk of missing safeguarding cues. Participants highlighted challenges around registering and accessing healthcare due to the physical closure of surgeries. Participants reported indirect discrimination, language and communication barriers, and lack of access to targeted and tailored COVID-19 information or interventions. In addition, migrants reported a range of specific beliefs around COVID-19 and on potential COVID-19 vaccines, from acceptance to mistrust, often influenced by misinformation. PCPs raised concerns that migrants may have increased risk factors for poor general health and to severe illness from COVID-19, in part due to their social and economic situation. Innovative opportunities were suggested to engage migrant groups through translated digital health advice using text templates and YouTube which merit further exploration.

**Conclusion:** Pandemic-related changes in primary care delivery may be here to stay, and some migrant groups are at risk of digital exclusion and may need targeted additional support to access services. As primary care networks operationalise the delivery of the COVID-19 vaccine, these findings provide critical information on specific strategies required to support migrant population to access primary care and overcome misinformation around COVID-19 and the COVID-19 vaccine.

**How this fits in:** The impact of pandemic-related shifts in primary care delivery on marginalised migrant groups, who may already face major disparities in accessing primary care, is poorly elucidated. We found that the rapid digitalisation of primary care services and physical closure of surgeries during the pandemic have amplified disparities in access to healthcare for specific migrant groups, with many lacking access to and capacity to use technology, compounded by language barriers. Migrants may be at increased risk of misinformation about COVID-19, which merits further consideration as COVID-19 vaccine roll out begins. Improved outreach to local migrant community organisations and places of worship, alongside co-designing with migrants more inclusive delivery approaches and creative integration of migrant ambassadors into information-sharing campaigns are needed. Primary care can maximise the opportunities of digitalisation for migrants through flexible engagement by multiple modalities (e.g. text, email, letter and YouTube videos) to provide targeted, translated advice and information, virtual group consultations for patients with a specific condition, and working with local leaders and NGOs to access and disseminate information through informal communication channels.

## Introduction

Migrants to the UK – particularly more recent arrivals and marginalised groups including refugees and asylum seekers – may face a multitude of barriers to accessing primary care, resulting in disparities in access to NHS care (1). Barriers include confusion around the NHS system and how to navigate it, language difficulties and discrimination, and, for some migrant groups, restricted entitlement to healthcare due to their immigration status (2). Migrants are a highly diverse group, but are considered to be disproportionately impacted by infectious diseases compared to the host population and may be under-immunised, with implications for health systems on arrival (3). There are concerns that these existing inequities have been exacerbated in the context of the COVID-19 pandemic, with emerging data highlighting the differential impact of SARS CoV-2 on migrant groups specifically, but also the wider ethnic minority population (including Black, Asian, and Minority Ethnic [BAME] groups) (4–6).

Increasing digitalisation of primary care has been a key feature of the pandemic, with the UK and other countries switching rapidly away from face-to-face consultations during the pandemic to virtual consultations via telephone, video, and online form-based communications (such as E-consult) and text communications (such as accuRx) (7,8). Digitalisation, however, can disproportionately disadvantage marginalised groups, amplifying existing structural inequalities through differential access and digital literacy, differential capacity to benefit, and differential motivation for use (9,10). In a recent report, Doctors of the World UK reported that migrants living in vulnerable circumstances in England have faced increased difficulties during the pandemic, including an inability to register with a GP, with digital exclusion and language barriers impacting on their ability to get advice and help with COVID-19 symptoms (11). These barriers may have important implications for COVID-19 vaccine roll-out, with Public Health England suggesting more flexible delivery models may be needed to reduce inequalities in vaccine uptake through primary care in at-risk BAME groups, (12) which includes some migrant populations.

In this study we aimed to seek the views of a broad range of primary care professionals (PCPs) and recently arrived migrants to the UK to explore and assess the specific impact of the pandemic on migrants and their access to primary care, implications for COVID-19 vaccination uptake, and to better understand potential facilitators and solutions to inform the immediate public health response.

## Methodology

### Study Design

We completed an in-depth qualitative study of both recently-arrived migrants (residing in the UK<10 years) and a broad range of primary care professionals (PCPs). This semi-structured interview study involved three iterative, linked phases. Phase 1 consisted of interviews with clinical PCPs and informed data collection and analysis for phase 2 (interviews with administrative PCPs) and phase 3 (recently-arrived migrants: defined as foreign-born; >18 years of age; <10 years in the UK). Topic guides were developed by the research team comprising FK and JC (GPs) and AD, SH, AC, SEH (academic researchers), and piloted with two GPs, with input from our Project Board of migrant representatives. Separate topic guides were developed for each phase. PCPs were asked broadly about their experience of providing healthcare and support to migrants, their views on the impact of the COVID-19 pandemic on migrant patients, and implications for COVID-19 vaccine roll-out. We sought views from migrants around their experiences during the pandemic, the impact of the COVID-19 pandemic on access to primary healthcare, and their views and concerns on the COVID-19 vaccine and delivery within their communities.

### Sampling and Recruitment

In Phase 1 and 2, PCPs were recruited using purposive sampling across urban, suburban, and rural settings. Further participants were identified by means of snowballing, with those recruited asked to contact colleagues about participating in the study. Recruitment packs including participant information sheets were disseminated through local Clinical Research Networks and advertised on GP bulletins, newsletters and through practice manager mailing lists.

In Phase 3, migrants were recruited using convenience and snowball sampling. Adverts for the study and participant information sheets were circulated to UK-focused migrant support groups and on social media, and shared with charities providing healthcare-related support specifically for migrant populations. Verbal (by phone) explanation of the study with an interpreter was offered to individuals interested in taking part.

### Ethics and informed consent

Ethics was granted by St George’s, University of London Research Ethics Committee (REC 2020.0058 and 2020.00630) and The Health Research Authority (REC 20/HRA/1674). For all 3 Phases participant information sheets were circulated, and signed informed consent was acquired prior to arranging a telephone interview. Participants gave consent to audio record interviews.

### Data Collection and Analysis

In-depth semi-structured interviews were conducted by telephone (by JC, FK, AD, AC, SEH) and lasted 30-90 minutes. We used this approach because it enables a targeted line of questioning, but also provides the opportunity for participants to explore in more detail areas that are important to them. Participants were compensated with an online shopping voucher (£20 for PCPs, £37 for migrants, due to a longer interview process). Each interview was audio-recorded then transcribed verbatim; transcripts were checked for accuracy and anonymised. Data collection ended when data saturation was reached, and no novel concepts were arising (13). Data were analysed inductively, informed by thematic analysis, which enables the researcher to explore collective experiences and understanding across a dataset through iterative analysis (14), using the stages set out by Braun and Clarke (2006). FK and JC undertook immersion to enable familiarisation and begin abstraction (15). Transcripts were analysed using NVIVO 12 and a comprehensive lists of codes was developed by FK and agreed by JC, AD and SH with any disagreements being resolved through negotiated consensus; key themes were conceptualised through further discussion with the wider research team and project board including migrant representatives.

## Results

We did 81 interviews in total (18^th^ June to 30^th^ November 2020). In Phase 1 we did 47 individual interviews with clinical PCPs including 25 General Practitioners (GPs), 15 practice nurses (PNs), 6 healthcare assistants (HCAs) and 1 clinical pharmacist. In Phase 2 we did interviews with 16 administrative staff including 11 practice managers and 5 receptionists/ other administrative staff. Characteristics of the PCPs interviewed are outlined in Table 1 (mean age 45 years; 84.4% female; range of ethnicities including White British, Indian, Pakistani). A wide range of practice sizes was represented with the majority being in an urban location (78.1%)

**Table 1:** Characteristics of Primary Care Professionals Interviewed.

|  | All participants | General Practitioner | Practice Nurse | Practice Manager/ Administration Team | Health Care Assistant / Clinical Pharmacist |
| --- | --- | --- | --- | --- | --- |
| **Number of participants n (%)** | 64 | 25 (39.1%) | 15 (23.4%) | 16 (25.0%) | 8 (12.5%) |
| **Age (mean)** | 45 (SD 11.8) | 44 (SD11.0) | 45 (SD11.9) | 48 (SD 11.6) | 41 (SD 12.7) |
| **Sex n (%)** |  |  |  |  |  |
| Female | 54 (84.4%) | 17 (26.6%) | 15 (23.4%) | 14 (21.9%) | 8 (12.5%) |
| Male | 10 (15.6%) | 8 (12.5) | 0 | 2 (3.1%) | 0 |
| **Ethnicity n (%)** |  |  |  |  |  |
| African | 4 (6.2%) | 0 | 3 (4.7%) | 0 | 1 (1.6%) |
| Other Asian background | 2 (3.1%) | 1 (1.6%) | 1 (1.6%) | 0 | 0 |
| Other mixed background | 3 (4.7%) | 1 (1.6%) | 0 | 1 (1.6%) | 1 (1.6%) |
| Other white background | 5 (7.8%) | 1 (1.6%) | 2 (3.1%) | 1 (1.6%) | 1 (1.6%) |
| Caribbean | 1 (1.6%) | 0 | 0 | 0 | 1 (1.6%) |
| Indian | 11 (17.2%) | 8 (12.5%) | 0 | 3 (4.7%) | 0 |
| Pakistani | 3 (4.7%) | 2 (3.1%) | 0 | 1 (1.6%) | 0 |
| White British | 32 (50.0%) | 12 (18.8%) | 9 (14.1%) | 9 (14.1%) | 2 (3.1%) |
| White Irish | 3 (4.7%) | 0 | 0 | 1 (1.6%) | 2 (3.1%) |
| **Practice Size** |  |  |  |  |  |
| <5000 | 6 (9.4%) | 1 (1.6%) | 1 (1.6%) | 2 (3.1%) | 2 (3.1%) |
| 5000-10,000 | 24 (37.5%) | 10 (15.6%) | 2 (3.1%) | 9 (14.1%) | 3 (4.7%) |
| 10,000-15,000 | 10 (15.6%) | 6 (9.4%) | 2 (3.1%) | 0 | 2 (3.1%) |
| 15,000-20,000 | 15 (23.4%) | 6 (9.4%) | 4 (6.2%) | 4 (6.2%) | 1 (1.6%) |
| >20,000 | 9 (14.1%) | 2 (3.1%) | 6 (9.4%) | 1 (1.6%) | 0 |
| **Practice Location** |  |  |  |  |  |
| Rural | 1 (1.6%) | 1 (1.6%) | 0 | 0 | 0 |
| Suburb | 13 (20.3%) | 7 (10.9%) | 2 (3.1%) | 2 (3.1%) | 2 (3.1%) |
| Urban | 50 (78.1%) | 17 (26.6%) | 13 (20.3%) | 14 (21.9%) | 6 (9.4) |

During Phase 3, we did 17 interviews with migrants, including 15 (88%) asylum seekers and 2 refugees (64% female; mean age 38 years (range 22-59 years); mean time in the UK 4 years (range 9 months-9 years). Participants originated from 14 countries across 5 WHO Regions (see Table 2).

**Table 2:**
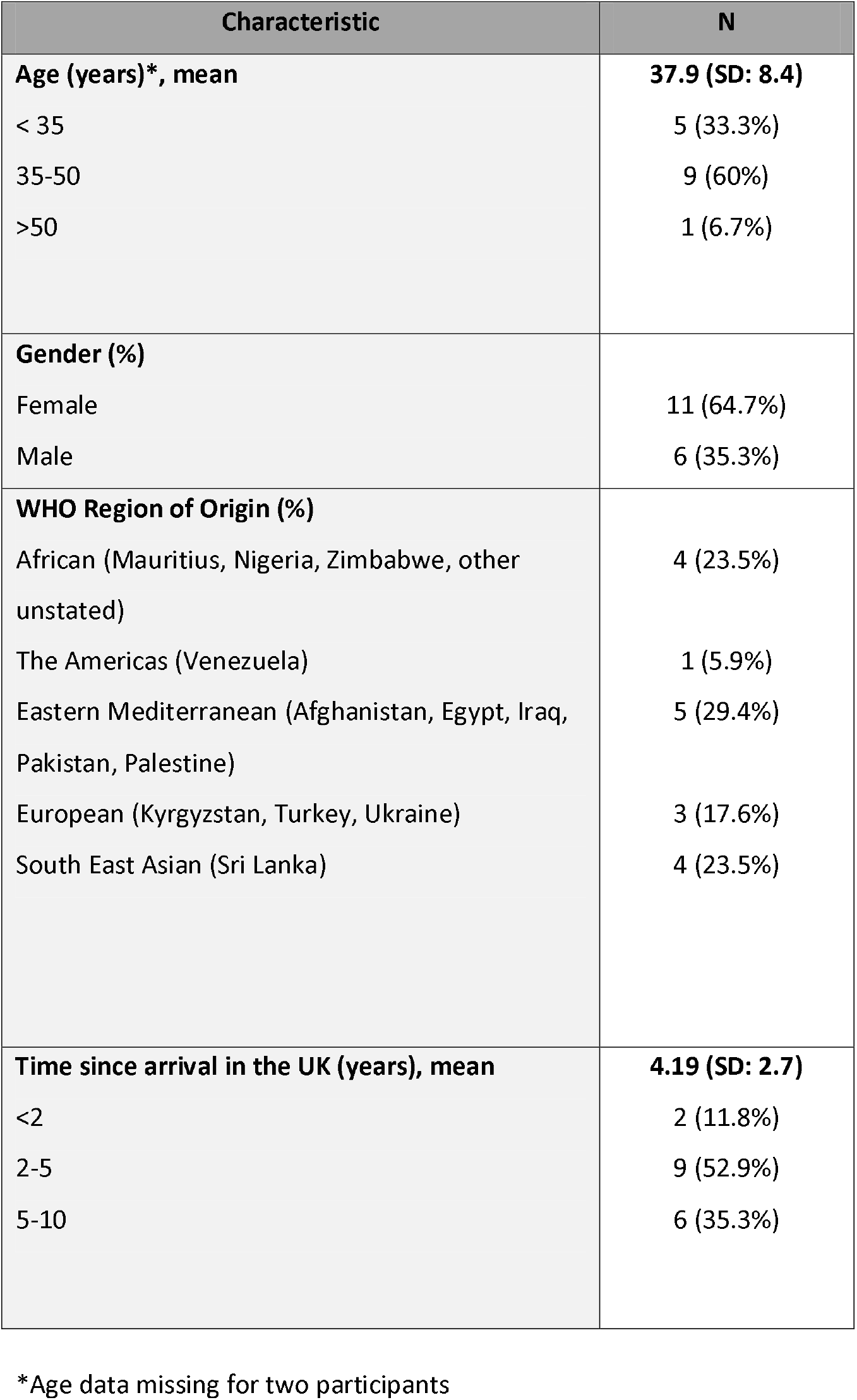
Characteristics of Migrants Interviewed.

### Impacts of the COVID-19 pandemic on migrants’ access to primary care

There was significant convergence on themes between migrants and all professional groups, who reported a range of impacts of the pandemics, risk factors for contracting SARS-CoV-2 and to COVID-19 vaccine roll-out, but also a range of opportunities and solutions which are summarised below.

#### Implications of digitalisation for migrant groups

PCPs described a shift to digitalisation of registration, appointments (moving from face-to-face to virtual consultations including telephone, video and online form-based), and giving of health information, prescriptions and “fit notes” (previously known as sick notes) by text, and were largely of the opinion that increased digitalisation was ‘here to stay’. Concerns were raised by many PCPs that lack of technology and challenges using it act as a barrier to access, although some did not feel this was a concern. Many migrants reported problems arising from digitalisation, citing lack of ownership of technological devices such as a computer or printer, or not knowing how to use, or being unable to afford to maintain them. Some PCPs reported an overall reduction in migrant registrations at their surgery, or decreased attendances by migrant groups, and that they perceived increased fear of entering the practice due to COVID-19 in these communities, and a preference for use of home remedies. Others perceived that this rapid digitalisation had increased access for patients who were generally fit and well, whilst exacerbating exclusion of more marginalised patients such as migrants.

*MIGRANT 9 ‘They ask you to go onto the website, fill out the form, sign it, scan it, and then send it back to them, so they can register you. I mean, I don’t have a scanner, I don’t have printers, then how can I kind of download it, scan? Or, if I can do it online, like an electronic signature, most people don’t know how to apply that. You need a computer. You can’t do that on your phone. So, those forms, for example, are not accessible at all for many people.’*

*ADMIN 1 ‘Everybody on this earth, unless you happen to be computer-phobic, knows how to use a smartphone…They probably have Facebook, they probably have Twitter, Instagram. If they can use that then our technology to book an appointment is very simple in comparison.’*

*GP 8 ‘One of the things that’s being pushed forward is remote consulting through e-Consult, for example… This is great for addressing need. Instead of actually addressing actual needs, which is different to perceived need, if that makes sense…. If you build more roads, you increase the traffic. So you’re not actually dealing with the demand in people who actually need the care. So they haven’t thought this through very well. And a lot of migrant populations are absolutely fine with technology. But, again, the outreach of that technology, or how to access it, isn’t known to them. And I feel the technology thing in the pandemic is going to widen [inequality of access].’*

*HCA 6 ‘And particularly, migrant patients did not want to come in. They tended to stay in the household…I just think, maybe they feel that at home, they’re safe…. They’re with their families, they do home remedies.’*

Other PCPs, particularly practice managers, commented that the increased use of technology has presented actual and potential creative solutions for marginalised migrant groups, such as using text messaging and being able to translate texts into the patient’s language, as well as targeted digital communications to support and encourage access, group video consultations, and the use of YouTube videos for delivering health advice. These solutions identified by participants are summarised in *Table 4*.

*GP16 ‘Yes. I’ve been texting in Turkish. I answer all my Turkish patients and I just know enough Turkish to check I’m not saying complete gobbledegook with Google Translate.’*

*ADMIN 8 ‘We do an awful lot of stuff by text, as we find that works really well and across language barriers… and migrant people really locked onto [YouTube] because you can see. It just works.’*

*HCA 6: ‘We sent out some text messages, just saying, we’re here, we are open, please come and see us… One of our receptionists is making phone calls to where we had very vulnerable migrant families. And just let them know that we could be here. We could visit them at home. We also made some COVID leaflets out as well, that was in Somali language and different languages, which we then posted to the vulnerable families that we have.’*

#### Social and Economic Factors Impacting Health Status and Access to Services

Concerns were cited by both PCPs and migrants that migrant groups had been disproportionately more likely to experience pandemic-related financial insecurity and that some may have faced increased exposure to SARS-CoV-2 due to their front-facing jobs. Migrant participants often expressed that financial concerns, social exclusion and poor living conditions, had a substantial negative impact on their mental health. Some clinical PCPs were optimistic that these impacts could be mitigated by a renewed health policy emphasis on supporting access for marginalised groups during the pandemic.

*MIGRANT 14 ‘It’s [the pandemic and lockdown] just making the situation for people worse, in a way, because people will start having suicidal thoughts, starting to think about the country that you came here from, there’s war, there’s poverty. It also makes people start to have conflict in the hotels, in the hostels, because people will be too stressed, they are thinking of things back home’.*

*PN13 ‘I think one of the challenges is the fact that, because of COVID, there’s going to be a lot more job insecurity for these people, which is going to have more of an impact on their healthcare anyway. So I think we’re going to see a lot of problems in terms of poverty and food banks and people who’ve got no recourse to public funds going into even more poverty.’*

Migrant participants highlighted being moved into cramped hotel or hostel accommodation, and raised concerns around additional costs resulting from the pandemic, for example needing to buy soap and masks when they are on very low budgets. They also reported a loss of access to support networks and community organisations during the pandemic, services that previously helped them to access healthcare and navigate the healthcare system.

*MIGRANT 17 ‘We live on £5, £6 daily, so on top of this you have to buy soap and you have to buy disinfectant, you have to buy a mask, it adds a lot of pressure on your budget’.*

*MIGRANT 18 ‘Before the pandemic, you know, people who are British volunteers used to help us, speak to the refugees to apply for those [help with healthcare costs]. But now everything is closed. But yes, people are still sick’.*

#### Language barriers compounding access issues

Language barriers were repeatedly reported by migrants and PCPs alike as a key challenge to migrants accessing primary care during the pandemic. Prior to the pandemic, a range of language support modalities were used in primary care, but several practices reported regularly relying upon translation by family members or Google Translate in the context of reduced funding for interpreters. Language barriers were perceived to have been increased due to digitalisation (e.g. surgeries closed necessitating reliance on telephone and video consultations; forms online only available in English). Some migrants reported that pandemic-related lockdowns had reduced access to friends or organisations that had previously translated for them during and outside of healthcare appointments, and had had a negative impact on their ability to understand health information, appointment letters, and messaging around COVID-19.

*GP15 ‘I think a face-to-face consultation between a recently arrived migrant, particularly the language barrier, is really, really difficult. And I think the phone conversations that I’ve had [because of the pandemic], have been significantly more so, to the point that, more often than not, if there’s going to be a language barrier, and I think it’s a complex problem, I’ll just book people in for a face-to-face. But to get to that point, they have to have been able to actually access the appointment system, which is either online or on the phone.’*

*MIGRANT 4 ‘None of them speaks English so they were not aware about the restriction and what is the rules. So I advised them and I told them what they should do, how they should act: Wearing face masks, coming back, washing their hands…if I don’t speak English and I get a letter, I could travel to go seek help from my friends and translate for me. But what if I don’t speak English and there is a lockdown where I cannot go out?’*

Some GPs expressed a lack of knowledge or desire to engage with digital consultations involving an interpreter, whilst several GPs and practice nurses expressed concerns about confidentiality and their ability to pick up on cues and safeguarding issues in digital consultations.

*GP3 ‘I think if there are language barriers, then absolutely [telephone consultations cause challenges for migrants]…I think it depends hugely on that, because I’ve not tried to have a conversation with an interpreter using this [telephone consultation]. But I imagine that must be quite challenging because you’ve got to sort out how to do that three-way. I don’t know.’*

*GP18 ‘I mean confidentiality is another issue in terms of people’s living situation and overcrowding and maybe they’re sharing computers or obviously rooms and phone calls. We don’t know who’s in the background when we ring people… there are quite a lot of safeguarding issues where it would be good to either have occasional or often appointments with people on face-to-face or at least being in their own space, with us, the private space.’*

However, some PCPs reported improved ability to organise language support and improved access through digital consultations.

*GP 1 ‘The E-consultation method has, surprisingly, shown how the migrant contacts with the surgery have actually increased, compared to pre-COVID.… And increased the reach towards patients who might have language barriers, because they have the ability now to take their time. Maybe use a translator when they’re writing, and write down their concerns. And we’ve got many patient examples, who we may have thought would have potentially struggled with accessing remote healthcare, because English was not their first language’*

#### Trust, Authority and Information

Both migrants and clinical PCPs, particularly HCAs, commented that both before and during the pandemic there has been a lack of information targeted and tailed towards migrants about access to healthcare services and basic information relating to the virus itself combined with an ineffective, passive approach towards distributing such information where it does exist. Migrants reported a lack of understanding of health service changes and that there is considerable misinformation circulating among migrant communities about COVID-19, and the COVID-19 vaccine.

*MIGRANT 6 ‘Some of the people, culturally, they don’t believe that such a virus exists. They think that it’s 5G or something else. They rely on other news, so, that’s why, in order to change their minds and kind of make them believe, there should be an effective system of information’.*

*MIGRANT 15 ‘And we had several issues which were urgent, e.g., my husband, he couldn’t move, he had a pain in his back. But we felt we can’t go to the GP because of this COVID. My son complained for some time about his [tooth] pain I thought we can go to the dentist? We can go to the GP? I worried. I thought, what if it’s very, very urgent, what do we do? Even now, I don’t know. If something’s urgent, is the emergency department working at the moment?’*

*GP8 ‘[Regarding information-giving around COVID-19] It’s been very blustery from politicians. If English wasn’t your language and you watched a press conference, it’s quite hard to work out what is going on actually. And then the public health messaging, again, it’s not always been very simplistic. It has been changing. It’s very difficult. It’s only because organisations like Doctors of the World, etc… The big issue, basically, is language getting out there to people who need it.’*

Both migrants and PCPs stated a belief that some migrant patients have low levels of health literacy and do not believe in, or trust, science, the UK health system or government, and tend to seek religious or peer input into their decision-making. Concerns were raised around the links between immigration and access to health care. These alternative sources of information that some migrants may rely on, and lack of trust in UK-based authorities, were considered to have created particular confusion and mistrust during the pandemic among migrant communities.

*MIGRANT 8 ‘They [social media groups] were spreading a lot of information like don’t go outside tonight because the government will be spreading the powder that will stop COVID. And the funny thing is people believe it because somebody sent them…Like I see in the Russian-speaking group on Facebook so much confusion, so much misunderstanding of the system…I think this is where people make decisions. They will not trust a GP. Even after 16 years in the country.’*

*GP1 ‘I think they follow advice, and healthcare advice, not necessarily from doctors but from, let’s say, elders within their family society, local community places of worship.’*

Several migrants and clinical PCPs stated that face-to-face appointments are critical for PCPs to build trust with migrant patients and that the pandemic has made it much harder to build this trust through virtual consultations.

*GP22 ‘I find that if you do spend a bit more time with people right at the beginning, on what the issues are… that then makes future consultations much more straightforward. Because you’ve done the relationship-building bit of that, and I think that’s much harder to do via telephone, remotely, than it is face-to-face.’*

#### Indirect discrimination against migrants through a ‘one size fits all’ approach

Several migrants suggested that pandemic-related changes in primary care have utilised a ‘one size fits all’ approach, and flexibility is essential to ensure effective and equitable access healthcare. Practices’ approach to digitalisation often failed to consider that some groups might struggle to access or use technology, and took a rigid approach to not seeing patients face-to-face even in situations where communication challenges or lack of interpreters would otherwise significantly affect the quality of the consultation.

*MIGRANT 4 ‘They should not use just one way of contact which is like via the phone …please find some way to help. Rather than just putting the blame on that patient that we are, you are missed …Not everybody has the same opportunity or access.’*

*MIGRANT 1 ‘I think that would be better if they would have a little bit GPs open so we could talk with them because normal GPs have access to the interpreters, so they can manage to talk with them and see what our needs are sometimes. But it was completely shut down [during the pandemic] which is also a terrible thing to do.’*

Both migrants and PCPs recognised that the physical closure of surgeries during the pandemic led to indirect discrimination (in which a universal policy disproportionately affects specific groups), because migrants have lost practical support from receptionists who previously helped them to register and access appointments, and may no longer receive signposting, screening services and new patient health checks-interventions particularly benefitting migrant communities.

*GP18 ‘So although our registration seems easy, in COVID I expect it’s really difficult for people, because they can’t just walk in and get forms and do it in the waiting room. At least our receptionists speak a mixture of languages. They could help people fill in the forms. So that kind of thing isn’t happening so much.’*

*GP14 ‘We don’t get any intervention with them [migrants] at reception like we used to [due to the pandemic]. So, we’re not capturing any of the HIV testing or the chlamydia testing or the TB testing either.’*

#### Risk Factors for COVID-19 and COVID-19 Vaccination Roll-Out

PCPs and migrants alike reported concerns that pre-existing distrust of vaccinations and low health literacy in migrant communities – alongside widespread misinformation around the vaccine could negatively affect uptake of the COVID-19 vaccine in some migrant groups. Migrants reported a range of beliefs that COVID-19 is a ‘hoax’ or ‘Western disease’, fear of discrimination or being used as ‘guinea pigs’, and a reliance on ‘home remedies’. These issues are explored in *Table 3*.

**Table 3:** Mistrust and Misinformation Relating to the COVID-19 Vaccine Among Migrant Communities.

| Domain | Key Finding | Example Quote |
| --- | --- | --- |
| **Decision-making about whether to have the vaccine** | Mistrust of UK Doctors, government, or around immigration status | HCA 6 'We're going to have major issues, because I think there's, as I said before, there's a huge distrust around the government.' |
|  | Making decisions on advice from peers, social media or religious leaders | MIGRANT 8 '[On WhatsApp] they were spreading a lot of information like don't go outside tonight because the government will be spreading the powder that will stop COVID. And the funny thing is people believe it because somebody sent them.' |
|  | Contradictions between information sources and Indecision | MIGRANT 15 'Our church leaders, they're all saying to us not to be vaccinated. And, to be honest, I have no right correct answer, I'm confused. I'm not 100% sure that this is actually connected with demonic people who are trying, how can I say, to put their power on people.' |
| **Beliefs about COVID-19** | It is a hoax | MIGRANT 9 'Some of the people, culturally, they don't believe that such a virus exists. They think that it's 5G or something else.' |
|  | It is a 'Western disease' | PN13: 'In a lot of ethnic countries, COVID hasn't had the same impact as it has especially within Western Europe and America, and I think it's being seen as very much a European infection. It doesn't impact on BAME communities as much as they say.... There's racial connotations to it as well, but any vaccine that they bought will be similar to the AIDS infection in Africa and people bringing infections to new countries.' |
|  | Natural remedies will protect you | MIGRANT 3 'You just give one teaspoon honey with seven flaxseeds every morning just give them, just to protect from viral.' |
| **Beliefs about the COVID – 19 Vaccine** | Migrant communities have not been included in trials | MIGRANT 18 'The main problem is that we not having the same community participating too much in the trials' |
|  | Migrants will be used as guinea pigs | MIGRANT 9 'If I go there they might be using me as a guinea pig or I don't know. They might be using me for their own things. I don't trust them.' |
|  | Migrants will be discriminated against in receiving the vaccine | MIGRANT 16: 'We have that feeling we'd be the last to have the vaccination. Yes, we have that' |
|  | The vaccine will not work, or not be safe, or result in contracting COVID-19 | PN11 'They fear catching the vaccine and they would fear that it is a mass immunisation programme, that there could be lots of people around.' |
|  | The vaccine will control or microchip people | GP16: 'He said, a doctor? So glad, you can tell me. Is it true, the vaccine for this virus, that it's going to be microchip in it and track me around?' |

**Table 4:**
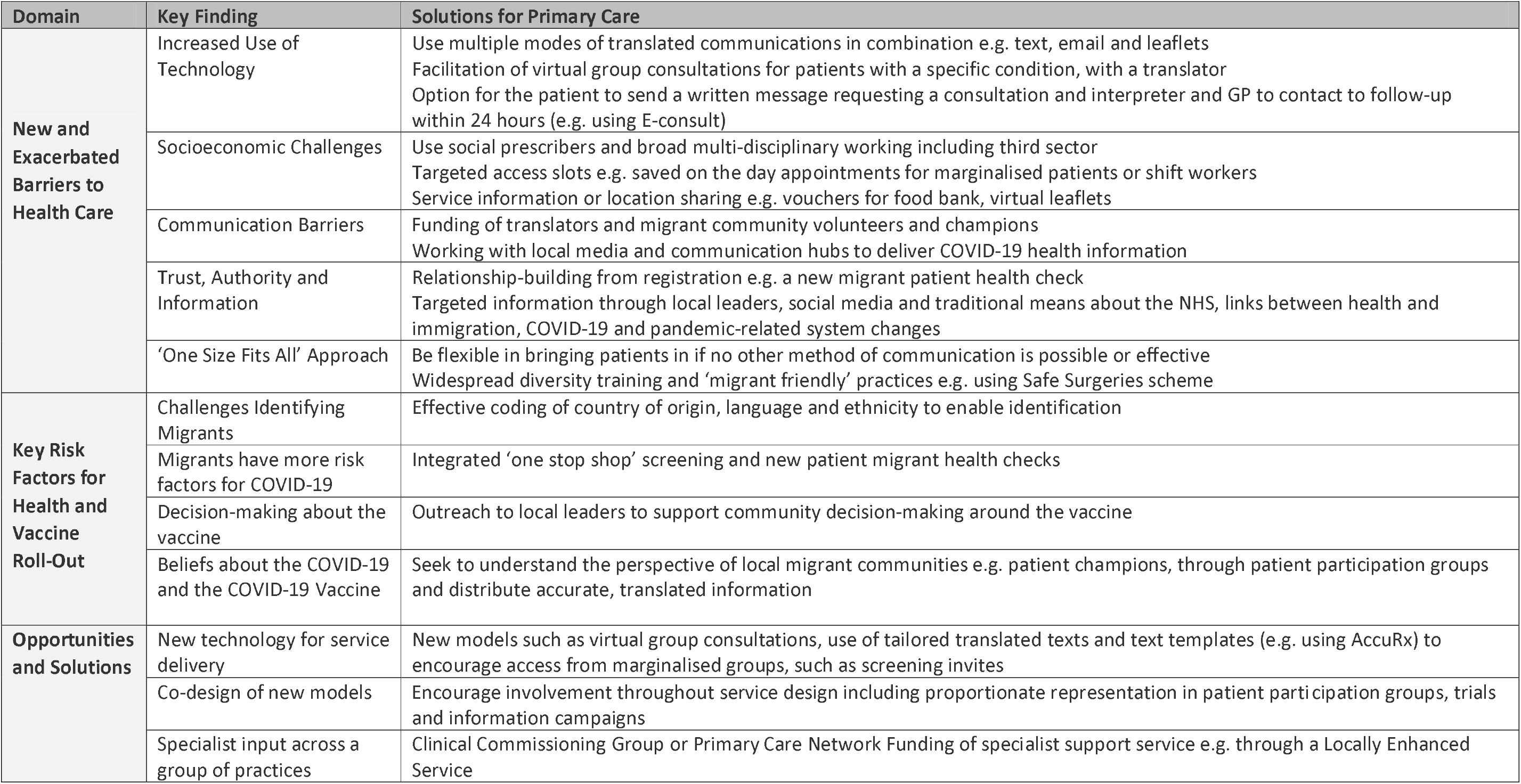
Key Findings and Solutions Identified.

| Domain | Key Finding | Solutions for Primary Care |
| --- | --- | --- |
| **New and Exacerbated Barriers to Health Care** | Increased Use of Technology | Use multiple modes of translated communications in combination e.g. text, email and leaflets<br>Facilitation of virtual group consultations for patients with a specific condition, with a translator<br>Option for the patient to send a written message requesting a consultation and interpreter and GP to contact to follow-up within 24 hours (e.g. using E-consult) |
|  | Socioeconomic Challenges | Use social prescribers and broad multi-disciplinary working including third sector<br>Targeted access slots e.g. saved on the day appointments for marginalised patients or shift workers<br>Service information or location sharing e.g. vouchers for food bank, virtual leaflets |
|  | Communication Barriers | Funding of translators and migrant community volunteers and champions<br>Working with local media and communication hubs to deliver COVID-19 health information |
|  | Trust, Authority and Information | Relationship-building from registration e.g. a new migrant patient health check<br>Targeted information through local leaders, social media and traditional means about the NHS, links between health and immigration, COVID-19 and pandemic-related system changes |
|  | ‘One Size Fits All’ Approach | Be flexible in bringing patients in if no other method of communication is possible or effective<br>Widespread diversity training and ‘migrant friendly’ practices e.g. using Safe Surgeries scheme |
| **Key Risk Factors for Health and Vaccine Roll-Out** | Challenges Identifying Migrants | Effective coding of country of origin, language and ethnicity to enable identification |
|  | Migrants have more risk factors for COVID-19 | Integrated ‘one stop shop’ screening and new patient migrant health checks |
|  | Decision-making about the vaccine | Outreach to local leaders to support community decision-making around the vaccine |
|  | Beliefs about the COVID-19 and the COVID-19 Vaccine | Seek to understand the perspective of local migrant communities e.g. patient champions, through patient participation groups and distribute accurate, translated information |
| **Opportunities and Solutions** | New technology for service delivery | New models such as virtual group consultations, use of tailored translated texts and text templates (e.g. using AccuRx) to encourage access from marginalised groups, such as screening invites |
|  | Co-design of new models | Encourage involvement throughout service design including proportionate representation in patient participation groups, trials and information campaigns |
|  | Specialist input across a group of practices | Clinical Commissioning Group or Primary Care Network Funding of specialist support service e.g. through a Locally Enhanced Service |

Clinical PCPs were concerned about the increased risk factors in migrants making them vulnerable to contracting and suffering serious illness from COVID-19, for example multi-morbidities, ethnicity and low socio-economic status.

*GP22 ‘The [migrant] population is a very high risk population, because of obesity and diabetes, ethnicity and other co-morbidities. And we have seen a lot of people dying in the community.’*

#### Opportunities and Solutions

PCPs expressed views that the new ways of working during the pandemic had also led to innovative solutions that could strengthen engagement with marginalised groups such as migrants, and has led to the generation of new ideas to inform service delivery beyond the pandemic. Creative solutions raised by participants included initiatives to provide targeted, translated health advice by text message and via YouTube videos, and new targeted community outreach approaches to faith and community leaders to access their communication networks and tackle misinformation about COVID-19 (*Table 4*). The delivery of new specialist services was reported, including specifically funded services working across multiple practices to coordinate interpreting services and community volunteers for specific marginalised groups including migrants. There was a consensus that to facilitate COVID-19 vaccine uptake in migrant groups, clear, concise and language-specific written and non-written resources needed to be developed, ideally centrally, and then made available for local or ‘hyperlocal’ distribution. In addition, participants noted that Clinical Commissioning Groups (CCGs), GP practices and pharmacies must pro-actively reach out to migrant communities and their institutions (religious and otherwise) to encourage uptake and work together to co-design solutions.

*GP16 ‘There could be some great accuRx [text messaging] templates for new migrant patients…Have you recently arrived in the UK? Would you like to get some health screening? Why not ask for a health screening appointment at the practice?’*

*GP8 ‘Who are the faith leaders of those communities, who runs them, and how are they communicating at the moment? Because we know, for example, if you look at people who are Muslim, they’re not going to pray together. So, therefore, that means we must know that the information they’re getting must be coming from their faith leaders. They must be having the call to prayer; they’re in their own homes. There must be a communication network to do that.’*

*PN13 ‘Because this is a targeted clinic, it meant that we were able to get volunteers from within the Mandarin and Cantonese-speaking communities. There was always a translator. There was always some leaflets and information as well in different languages. So I think that helps so much… volunteers who are part of the community as well. I think that builds trust within the healthcare professional and the patient.’*

*GP11 ‘And I don’t mean just written information [about the COVID-19 Vaccine], there needs to be more than that because, obviously, a lot of these people don’t actually read the language that they speak. So it has to be more sort of interpreted and through that to sort of, perhaps, work with the population, maybe with the religious leaders and so on to get people involved.’*

## Discussion

### Summary

We report a range of perspectives and experiences reported by both recently-arrived migrants and PCPs in the UK around the impact of COVID-19 on UK migrant communities and their access to healthcare, and their views around future COVID-19 vaccine roll out. We found that the rapid shift to digitalisation has exacerbated existing inequalities in access to healthcare for specific migrant groups through lack of access to or knowledge of technology with concerns expressed about language barriers, difficulties building trust and the risk of missing safeguarding cues virtually. The physical closure of some surgeries has led to challenges for migrants in registering and accessing primary care.

Communication barriers, feeling left behind, and lack of access to information were widely raised by migrants. Additionally, migrants reported specific views around COVID-19 and the associated vaccination, ranging from acceptance to fears that COVID-19 was a hoax and other misinformation, often originating from social media or word of mouth. During the pandemic, some migrants experienced increased risk factors to their health and severe illness from COVID-19, partially resulting from their economic and social situations. However, PCPs reported that pandemic-related changes to healthcare delivery may be here to stay, and have led to the development of several innovations in service delivery such as translated digital health advice using text templates and YouTube. *Table 4* provides a summary of the key findings and implications for primary care.

### Strengths and Limitations

Our study has generated valuable insights into the experiences of both migrants and PCPs during the COVID-19 pandemic. The findings are of direct immediate relevance to the ongoing public health response, in terms of supporting primary care providers with delivering services to marginalised migrant groups and in understanding the unique risk factors of this group. The scale of this study- the use of multiple phases, and the engagement of diverse voices including the migrants themselves and a range of different primary care professionals from across the country and multiple settings-enhanced validity. The similarities in perspectives between migrants and professionals was striking.

Next steps would be to engage migrants from all dominant nationality groups in the UK and different types of migrants (e.g. labour migrants, undocumented migrants) to explore the culture-specific impacts of COVID-19, and age-related differences within the different migrant populations, which could be particularly relevant to a COVID-19 vaccine targeting older groups. Additionally, the impact of the researchers’ ethnicity and social background, and professional training, may have influenced responses through perceived power differentials and social desirability bias, in a desire to express views they believe will be received favourably.

### Comparison with existing literature

Whilst there is a growing body of research exploring the impact of COVID-19 on BAME groups, the specific impact on migrant groups had not been explored in depth. Migrants may have a range of unique risk factors and vulnerabilities to COVID-19 (6) and face barriers to health care and poor health outcomes in the UK and Europe (3), which we found was exacerbated during the pandemic due to the rapid shift to digital healthcare provision. These issues are supported by research findings from Doctors of the World UK (11), who found that migrants experienced a lack of access to key COVID-19 public health messages, in part due to most health information about COVID-19 being published in English, a key theme in our study, which combined with low health literacy leaves these groups vulnerable to misinformation. This demonstrates the need for linguistically and culturally appropriate health information; the Swedish ‘corona lines’ – specialist phone services provided in various languages offering advice and COVID-19 triaging are one example of good practice (16).

Other studies pre-pandemic are conflicting, suggesting that there is a digital divide in resources needed to participate in remote consultations (7), and that minority groups such as migrants are more likely to miss virtual appointments (17), but can also benefit more than majority groups if interventions are specifically designed with their needs and capabilities in mind (18). Leite, Hodgkinson & Gruber (2020) have highlighted that if barriers such as access to broadband, digital literacy, and privacy and protection of patient data can be overcome, then digitalisation provides a crucial way to ease the impact of the pandemic, from enabling virtual triage to mitigating the negative psychological effects of social isolation (19). This concurs with our study, which found that where migrants could access and use technology, the use of translated texts and YouTube to provide health information and support access – as well as virtual group consultations with language support, might prove innovative solutions to improving access beyond the pandemic.

We suggest solutions to improve access to primary care for migrants in this pandemic, demonstrating the need for flexible and adapted policies in order to minimise disparities (20). Interventions to address structural inequalities and social and economic constraints that reduce migrants’ autonomy to be able to protect themselves from suffering serious illness from COVID-19 are needed (4,11) and to ensure culturally and linguistically appropriate information about COVID-19 and the COVID-19 vaccine (21).

Our findings provide insight into factors likely to impact on COVID-19 vaccine roll-out in migrant communities, concurring with previous studies that demonstrate that migrants may trust their social networks over medical professionals (22) and that they may be more likely than the general population to believe COVID-19 misinformation (23) and mistrust COVID-19 vaccine research (21). Specifically engaging diverse migrant groups in the UK, and co-designing interventions to facilitate COVID-19 vaccine uptake, is therefore a crucial next step.

### Implications for practice and research

Practices should seek to ensure they can identify migrants within their populations, that they understand their needs through proactive engagement with local community organisations, and that they are providing language-specific advice about COVID-19 and changes in service provision in the pandemic through multiple modalities (e.g. text, email, letter and posters in local community hubs). As primary care networks operationalise the delivery of the COVID-19 vaccines, our findings provide critical information about how primary care providers can meet the needs of migrant patients (*Table 4*). This includes considering co-design of delivery approaches, ensuring availability of interpreters and translated culturally-appropriate vaccine advice, alongside creative integration of migrant ambassadors into vaccine centres, and information-sharing campaigns. Further research should seek to compare and evaluate the success of different virtual consultation approaches in this group, and the success of targeted and community-based or co-designed service delivery models in improving access to healthcare and strengthening COVID-19 vaccine uptake.

## Ethical approval

Ethics was granted by St George’s, University of London Research Ethics Committee (REC 2020.0058 and 2020.00630) and The Health Research Authority (REC 20/HRA/1674).

## Data Availability

Data are available on request from the authors.

## Acknowledgements

We thank all participants for sharing their views and experiences. This study is funded by the NIHR and the Academy of Medical Sciences. FK is supported by a Health Education England / National Institute for Health Research (NIHR) Academic Clinical Fellowship. JC is funded by a National Institute for Health Research (NIHR) in-practice clinical fellowship (NIHR300290). The views expressed are those of the author(s) and not necessarily those of the NHS, the NIHR or the Department of Health. AD and SEH are supported by Medical Research Council PhD studentships (MR/N013638/1). SH is funded by the NIHR (NIHR Advanced Fellowship NIHR300072) and the Academy of Medical Sciences (SBF005\1111), and by the European Society of Clinical Microbiology and Infectious Diseases (ESCMID) through a joint ESCMID Study Group for Infections in Travellers and Migrants (ESGITM) and ESCMID Study Group for Mycobacterial Infections (ESGMYC) Research grant. AFC is funded by the Academy of Medical Sciences (SBF005\1111) and the NIHR (NIHR300072).

## References

1. Rafighi E, Poduval S, Legido-Quigley H, Howard N. National Health Service Principles as Experienced by Vulnerable London Migrants in ‘Austerity Britain’: A Qualitative Study of Rights, Entitlements, and Civil-Society Advocacy. Int J Health Policy Manag. 2016 May 8;5(10):589–97.

2. Kang C, Tomkow L, Farrington R. Access to primary health care for asylum seekers and refugees: a qualitative study of service user experiences in the UK. Br J Gen Pract. 2019 Aug 1;69(685):e537–45.

3. Noori T, Hargreaves S, Greenaway C, van der Werf M, Driedger M, Morton RL, et al. Strengthening screening for infectious diseases and vaccination among migrants in Europe: What is needed to close the implementation gaps? Travel Medicine and Infectious Disease. 2020 May 7;101715.

4. Patel P, Hiam L, Sowemimo A, Devakumar D, McKee M. Ethnicity and covid-19. BMJ. 2020 Jun 11;369:m2282.

5. Public Health England. Disparities in the risk and outcomes of COVID-19 [Internet]. 2020 [cited 2021 Jan 7] p. 92. Available from: https://assets.publishing.service.gov.uk/government/uploads/system/uploads/attachment_data/file/908434/Disparities_in_the_risk_and_outcomes_of_COVID_August_2020_update.pdf

6. Hayward SE, Deal A, Cheng C, Crawshaw AF, Orcutt M, Vandrevala TF, et al. Clinical outcomes and risk factors for COVID-19 among migrant populations in high-income countries: a systematic review. medRxiv [Preprint]. 2020 Dec 22;2020.12.21.20248475.

7. Ortega G, Rodriguez JA, Maurer LR, Witt EE, Perez N, Reich A, et al. Telemedicine, COVID-19, and disparities: Policy implications. Health Policy and Technology. 2020 Sep 1;9(3):368–71.

8. Bhaskar S, Bradley S, Chattu VK, Adisesh A, Nurtazina A, Kyrykbayeva S, et al. Telemedicine Across the Globe-Position Paper From the COVID-19 Pandemic Health System Resilience PROGRAM (REPROGRAM) International Consortium (Part 1). Front Public Health [Internet]. 2020 Oct 16 [cited 2021 Jan 7];8. Available from: https://www.ncbi.nlm.nih.gov/pmc/articles/PMC7596287/

9. National Academies of Sciences, Engineering, and Medicine, Health and Medicine Division, Board on Population Health and Public Health Practice, Committee on Community-Based Solutions to Promote Health Equity in the United States. Communities in Action: Pathways to Health Equity [Internet]. Baciu A, Negussie Y, Geller A, Weinstein JN, editors. Washington (DC): National Academies Press (US); 2017 [cited 2021 Jan 7]. Available from: http://www.ncbi.nlm.nih.gov/books/NBK425848/

10. Toyama K. Technology as amplifier in international development. In: Proceedings of the 2011 iConference on -iConference ‘11 [Internet]. Seattle, Washington: ACM Press; 2011 [cited 2021 Jan 7]. p. 75–82. Available from: http://portal.acm.org/citation.cfm?doid=1940761.1940772

11. Doctors of the World. A Rapid Needs Assessment of Excluded People in England During the 2020 COVID-19 Pandemic [Internet]. London: Doctors of the World; 2020 [cited 2021 Jan 11] p. 101. Available from: http://www.doctorsoftheworld.org.uk/wp-content/uploads/2020/05/covid19-full-rna-report.pdf

12. Campos-Matos I, Mandal S. Annex A: COVID-19 vaccine and health inequalities: considerations for prioritisation and implementation [Internet]. London: Department of Health & Social Care; 2021 [cited 2021 Jan 7] p. 11. Available from: https://www.gov.uk/government/publications/priority-groups-for-coronavirus-covid-19-vaccination-advice-from-the-jcvi-30-december-2020/annex-a-covid-19-vaccine-and-health-inequalities-considerations-for-prioritisation-and-implementation

13. Saunders B, Sim J, Kingstone T, Baker S, Waterfield J, Bartlam B, et al. Saturation in qualitative research: exploring its conceptualization and operationalization. Qual Quant. 2018 Jul 1;52(4):1893–907.

14. Braun V, Clarke V. Thematic analysis. In: APA handbook of research methods in psychology, Vol 2:Research designs: Quantitative, qualitative, neuropsychological, and biological. Washington, DC, US: American Psychological Association; 2012. p. 57–71. (APA handbooks in psychology®).

15. Ritchie J, Spencer L. Qualitative data analysis for applied policy research [Internet]. Analyzing Qualitative Data. Routledge; 2002 [cited 2021 Jan 7]. Available from: https://www.taylorfrancis.com/chapters/qualitative-data-analysis-applied-policy-research-jane-ritchie-liz-spencer/10.4324/9780203413081-14

16. Valeriani G, Sarajlic Vukovic I, Lindegaard T, Felizia R, Mollica R, Andersson G. Addressing Healthcare Gaps in Sweden during the COVID-19 Outbreak: On Community Outreach and Empowering Ethnic Minority Groups in a Digitalized Context. Healthcare (Basel) [Internet]. 2020 Nov 1 [cited 2021 Jan 7];8(4). Available from: https://www.ncbi.nlm.nih.gov/pmc/articles/PMC7712425/

17. Kolb CM, Born K, Banker K, Barth PC, Aaronson NL. Improving Attendance and Patient Experiences During the Expansion of a Telehealth-Based Pediatric Otolaryngology Practice. Otolaryngol Head Neck Surg. 2020 Oct 20;0194599820965917.

18. Turnbull S, Cabral C, Hay A, Lucas PJ. Health Equity in the Effectiveness of Web-Based Health Interventions for the Self-Care of People With Chronic Health Conditions: Systematic Review. J Med Internet Res. 2020 Jun 5;22(6):e17849.

19. Leite H, Hodgkinson IR, Gruber T. New development: ‘Healing at a distance’—telemedicine and COVID-19. Public Money & Management. 2020 Aug 17;40(6):483–5.

20. Thorneloe R, Wilcockson H, Lamb M, Jordan CH, Arden M. Willingness to receive a COVID-19 vaccine among adults at high-risk of COVID-19: a UK-wide survey. PsyArXiv [Preprint] [Internet]. 2020 Jul 20 [cited 2021 Jan 7]; Available from: https://psyarxiv.com/fs9wk/

21. Ekezie W, Czyznikowska BM, Rohit S, Harrison J, Miah N, Campbell-Morris P, et al. The views of ethnic minority and vulnerable communities towards participation in COVID-19 vaccine trials. J Public Health (Oxf). 2020 Oct 30;

22. Udwan G, Leurs K, Alencar A. Digital Resilience Tactics of Syrian Refugees in the Netherlands: Social Media for Social Support, Health, and Identity. Social Media + Society. 2020 Apr 1;6(2):2056305120915587.

23. Loomba S, de Figueiredo A, Piatek SJ, de Graaf K, Larson HJ. Measuring the Impact of Exposure to COVID-19 Vaccine Misinformation on Vaccine Intent in the UK and US. medRxiv [Preprint] [Internet]. 2020 Oct 26 [cited 2021 Jan 7]; Available from: http://medrxiv.org/lookup/doi/10.1101/2020.10.22.20217513

